# Comparison of Different Diagnostic Criteria of ^68^Ga-Pentixafor PET/CT for the Classification of Primary Aldosteronism

**DOI:** 10.1101/2024.07.26.24311092

**Authors:** Xiangshuang Zhang, Furong He, Ying Song, Ying Jing, Jinbo Hu, Hang Shen, Aipin Zhang, Wenwen He, Zhengping Feng, Qifu Li, Hua Pang, Shumin Yang the Chongqing Primary Aldosteronism Study (CONPASS) Group

**Author notes:** Membership of the CONPASS Group is provided in the Acknowledgments. These authors contributed equally to this article as co-first authors (Xiangshuang Zhang, Furong He, Ying Song). Shumin Yang, Qifu Li and Hua Pang should be considered joint senior authors. **Reprint requests and correspondence:** Shumin Yang, Department of Endocrinology, The First Affiliated Hospital of Chongqing Medical University, No. 1 Youyi Street, Chongqing 400016, China. or Hua Pang, Department of Nuclear medicine, The First Affiliated Hospital of Chongqing Medical University, No. 1 Youyi Street, Chongqing 400016, China. or Qifu Li, Department of Endocrinology, The First Affiliated Hospital of Chongqing Medical University, No. 1 Youyi Street, Chongqing 400016, China.

## Abstract

**Background:** ^68^Ga-Pentixafor positron emission tomography/computed tomography (PET/CT) is an emerging method for the classifying primary aldosteronism (PA). How to use this method for PA classification is still controversial.

**Methods:** A retrospective study was conducted in patients with PA who underwent PET/CT. These patients had a classification diagnosis of unilateral PA (UPA) or bilateral PA (BPA) based on adrenal venous sampling or post-surgical outcomes. Area under the receiver operating characteristic curve (AUC), specificity and sensitivity were used to analyze the accuracy of the lateralization index (LI) based on adrenal maximum standardized uptake value (SUV_max_), dominant side SUV_max_ adjusted by liver, dominant side of SUV_max_ and visual analysis.

**Results:** A total of 208 PA patients were included, with 128 UPA and 80 BPA. The AUC for diagnosing UPA using LI and visual analysis were 0.82 [95% CI, 0·77-0·87] and 0.82 (95% CI, 0·76-0·87), respectively, higher than the dominant side of SUV_max_ [0.72, (95%CI, 0·65-0·78)] and dominant side SUV_max_ adjusted by liver [0.71,(95%CI, 0·64-0·77)]. Visual analysis showed a sensitivity of 0.73 (95%CI,0.65-0.81) and a specificity of 0.88(95%CI,0.80-0.95). The LI cutoff of 1.50 resulted the highest Youden Index of 0.59, with a sensitivity of 0.68 (95%CI,0.59-0.76) and a specificity of 0.91 (95%CI,0.83-0.96). When the LI cutoff was increased to 1.65, the sensitivity reduced to 0.61 (95%CI,0.53-0.70), while the specificity increased to 0.96 (95%CI,0.89-1.00).

**Conclusion:** Both LI and visual analysis of PET/CT could be used in the classification diagnosis of PA. Nevertheless, visual analysis is more sensitive, and LI is more advantageous in specificity.

## Introduction

Primary Aldosteronism (PA) is one of the most common causes of secondary hypertension, accounting for 5%-10% of all hypertensive patients^1-3^. The condition is characterized by the autonomous secretion of aldosterone due to cortical lesions, presenting clinically with hypertension and/or hypokalemia. Accurate differentiation between unilateral primary aldosteronism (UPA) and bilateral primary aldosteronism (BPA) is essential, as UPA typically requires surgical intervention, while BPA is managed with oral mineralocorticoid receptor antagonists like spironolactone^4^.

Currently, the primary methods for subtype diagnosis mainly rely on adrenal CT and adrenal venous sampling (AVS), however, the accuracy of CT is only 50%∼70%^5,6^. As a result, guidelines recommend AVS for PA patients, except for those with typical imaging and clinical features of aldosterone-producing adenoma (APA) who are younger than 35 and have a history of hypokalemia. Despite its necessity, AVS is invasive, costly, and technically challenging, highlighting the need for non-invasive diagnostic methods.

Our research has previously demonstrated the potential role of ^68^Ga-Pentixafor PET/CT in subtyping PA and using lateralization index (LI) based on adrenal maximum standardized uptake value (SUV_max_) resulted a 90% concordance rate with AVS^7^. However, other diagnostic criteria of ^68^Ga-Pentixafor PET/CT in subtyping PA, such as dominant side of SUV_max_, liver-adjusted SUV_max_ and visual analysis have also been reported^8-13^. As ^68^Ga-Pentixafor PET/CT is a newly emerging method in PA subtyping diagnosis, published studies were small sample-sized, with a minimum of nine and a maximum of 123 PA patients included, and in particular, the maximum sample size of BPA were only 33^8-13^.

This study aims to compare the accuracy of different diagnostic criteria of ^68^Ga-Pentixafor PET/CT in the classification diagnosis of PA by increasing the sample size and the result might provide useful information for clinical practice.

## Methods

### Study Design and Participants

This retrospective study was conducted at the First Affiliated Hospital of Chongqing Medical University in China from October 2021 to December 2023. Patients diagnosed with PA and who received ^68^Ga-Pentixafor PET/CT were included. The study protocol was approved by the Ethics Committee of the First Affiliated Hospital of Chongqing Medical University. Informed written consent was obtained from each participant.

Exclusion criteria were: (a) concurrent autonomous cortisol excess (cortisol after 1 mg dexamethasone suppression test ≥ 1.8 μg/dL)^14^; (b) incomplete data of ^68^Ga-Pentixafor PET/CT or AVS; (c) with inconclusive subtyping diagnosis (e.g. unsuccessful cannulation of adrenal veins, received adrenalectomy without AVS but lost to follow-up after surgery, achieved incomplete biochemical remission after partial adrenalectomy).

### Diagnosis of PA

The diagnosis of PA was previously described^7,15^. In brief, prior to screening, antihypertensive medications affecting the aldosterone-renin ratio (ARR) were discontinued or substituted with α-blockers or non-dihydropyridine calcium antagonists for 2-4 weeks. Patients with hypokalemia needed potassium supplementation to maintain serum potassium above 3.5 mmol/L as much as possible. Blood samples were collected in the morning after at least 2 hours of a seated position and a 15-minute rest period to measure plasma renin concentration (PRC) and plasma aldosterone concentration (PAC). The screening test was positive when the ARR was ≥ 20 pg·ml^−1^/μIU·mL^−1^ ^16,17^. Patients who tested positive underwent confirmatory testing using a captopril challenge test (CCT) and/or seated saline infusion test (SSIT). PA was confirmed if one of the following criteria was met: (a) PAC ≥ 110 pg/mL two hours after 50 mg captopril^18^; (b) PAC levels ≥ 80 pg/mL after 2L normal saline infusion^19^.

### 68Ga-Pentixafor PET/CT Scanning and Image Analysis

Patients diagnosed with PA did not require specific preparation for the PET/CT scan. The upper abdomen was scanned using a hybrid PET/CT scanner (Gemini TF 64, Philips) 10 and 40 minutes after intravenous tracer injection. Images were analyzed semi-quantitatively and visual analysis by two experienced nuclear medicine physicians. Adrenal CT findings were categorized into unilateral or bilateral lesions or bilaterally normal glands. Lesions included nodules and hyperplasia, with nodules defined as round or oval-shaped masses and hyperplasia is characterized by an adrenal thickness ≥10mm^20-22^.

Visual analysis: The morphology of adrenal lesions was assessed. The uptake level of adrenal lesions was compared to that of adjacent and contralateral adrenal tissue and lesions with high radioactive uptake were considered positive.

Semi-Quantitative Analysis: Regions of interest were identified as CT-observed lesions or those with suspected increased tracer uptake on PET. SUV_max_ was measured in these regions, and for adrenal glands without morphology changes, SUV_max_ was recorded. The liver’s average SUV_max_ of five 2 cm spheres served as the whole-body background. SUV_max_ at 10 minutes was used to calculate the lateralization index (LI) based on SUV_max_ and dominant side of SUV_max_ adjusted by liver (DSAL). The side with higher SUV_max_ in both adrenal glands is the dominant side. LI based on SUV_max_ was defined as (SUV_max_ of dominant side)/ (SUV_max_ of nondominant side), and DSAL as (SUV_max_ of dominant side in adrenal)/(SUV_max_ of liver)^7^.

### AVS

The AVS was performed as previously described^15^. In brief, patients underwent AVS in the morning between 08:00 and 12:00. Normal saline or ACTH was administered as continuous infusion which was started 30 minutes before AVS. Blood samples were collected sequentially from right and left adrenal veins. Three tubes of blood (2ml for one tube) in each adrenal vein were collected consecutively, and one tube of blood in the inferior vena cava (IVC) was collected immediately after the collection of each side of the adrenal vein blood. The average results of three adrenal vein blood samples were used for index calculation. Cortisol and aldosterone were measured in each sample. The selectivity index (SI) was defined as cortisol (adrenal vein)/cortisol (peripheral vein). Successful cannulation of AVS was determined by SI ≥ 2 in non-ACTH stimulated patients, while SI ≥ 3 in ACTH stimulated patients. LI was defined as [aldosterone-cortisol ratio of dominant adrenal vein)]/ [aldosterone-cortisol ratio of nondominant adrenal vein] while contralateral suppression was defined as [aldosterone-cortisol ratio of non-dominant adrenal vein] < [aldosterone-cortisol ratio of peripheral vein]. UPA diagnosis was made if LI ≥ 4 or LI 2-4 with contralateral suppression and typical adenoma on the dominant side by CT, while LI < 2 or LI 2-4 not meeting the above criteria indicated BPA^23,24^.

### Criteria for Classification

Definitive subtype diagnosis was based on AVS and/or post-adrenalectomy outcomes. For patients received adrenalectomy, based on PASO criteria^25^, patients achieving complete biochemical success were classified as UPA, while those not achieving complete biochemical success were categorized as BPA. Patients without surgery were classified according to AVS results.

### Statistical analysis

Statistical analyses were performed using SPSS Statistics Version 23.0 (IBM, Armonk, New York). Data distribution was assessed using the Kolmogorov-Smirnov test. Normally distributed variables were presented as mean and standard deviation (SD) and analyzed using Student’s t-test. Skewed distribution variables were presented as median (interquartile range) and analyzed using the Mann-Whitney U test. Categorical variables were described as percentages and analyzed using the χ2 test or Fisher’s Exact Test. The area under the curve (AUC) was calculated using MedCalc software 19.5.2. P values less than 0.05 were considered statistically significant.

## Results

### 1. Clinical characteristics of patients

In this study, 295 patients were included initially, but 87 were excluded, leaving 208 patients for final analysis. The cohort consisted of 128 patients diagnosed with UPA and 80 with BPA (Figure 1). There were no significant differences in age, gender, or blood pressure between the UPA and BPA groups (P > 0.05). However, the UPA group had a lower body mass index, serum potassium levels, and PRC compared with the BPA group. Conversely, PAC, PAC post-CCT, PAC post-SSIT were higher in the UPA group than in the BPA group (P<0.05) (Table 1).

**Figure 1.**
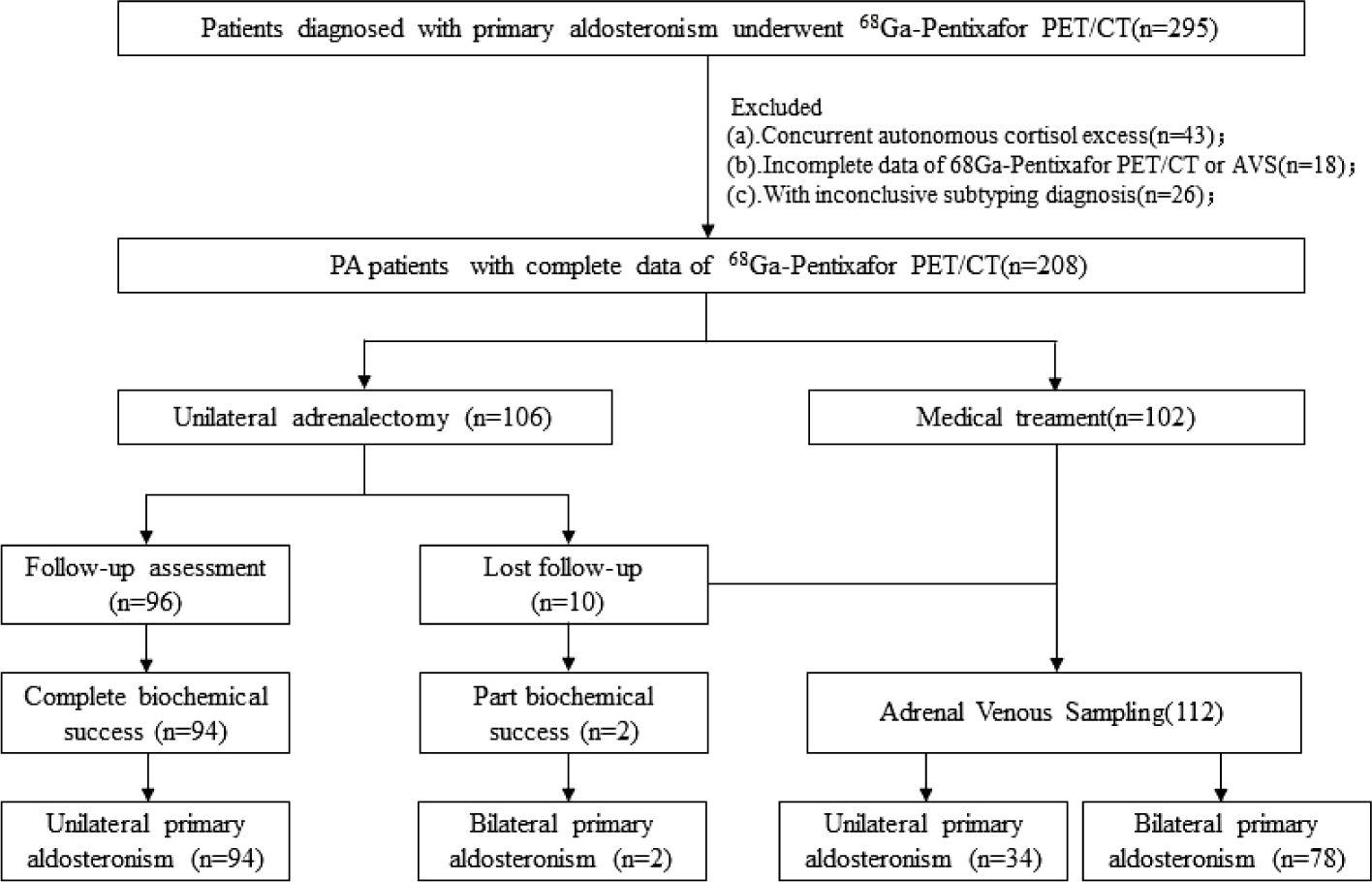
Flowchart of the Study.

**Table 1.**
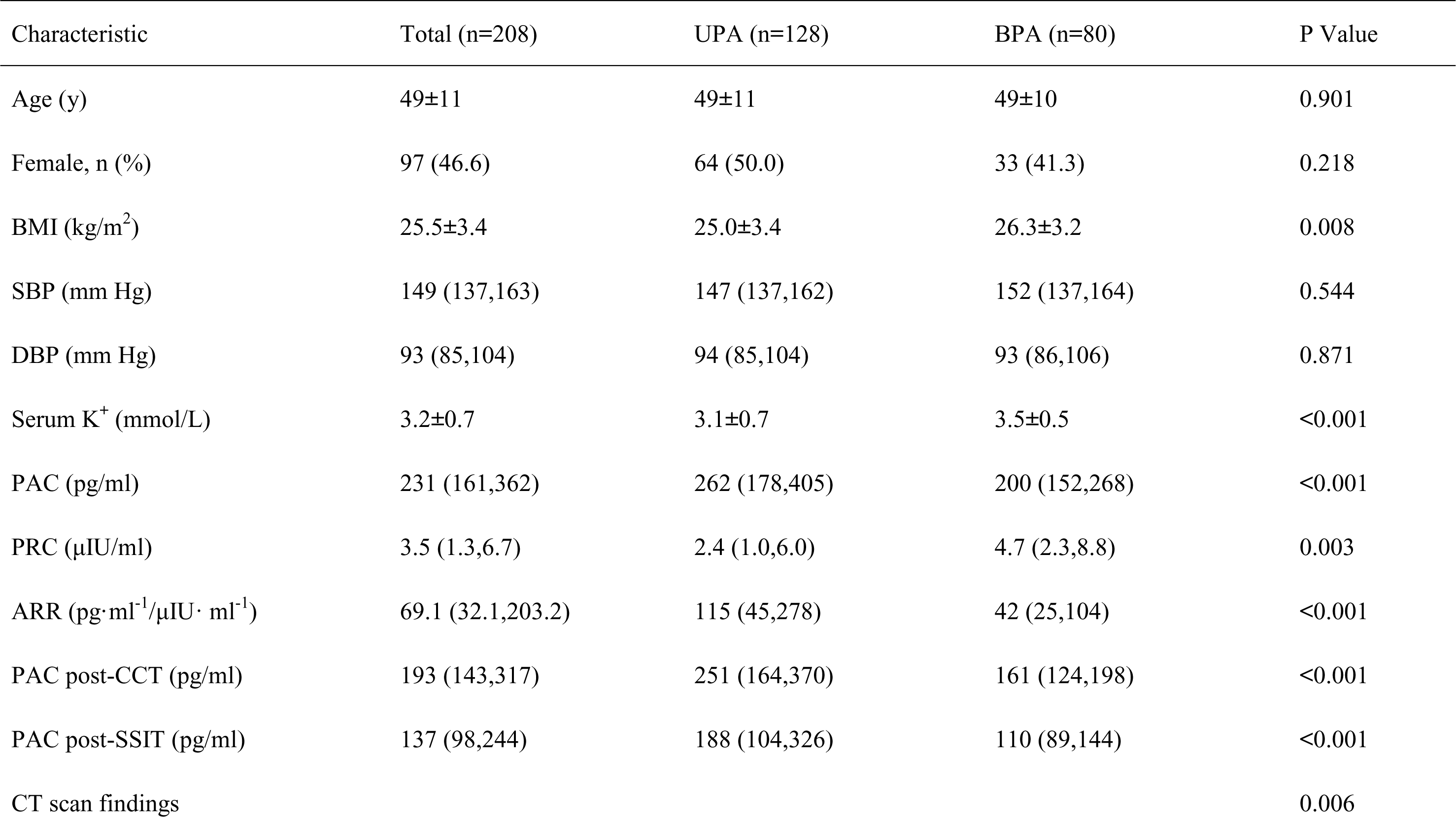
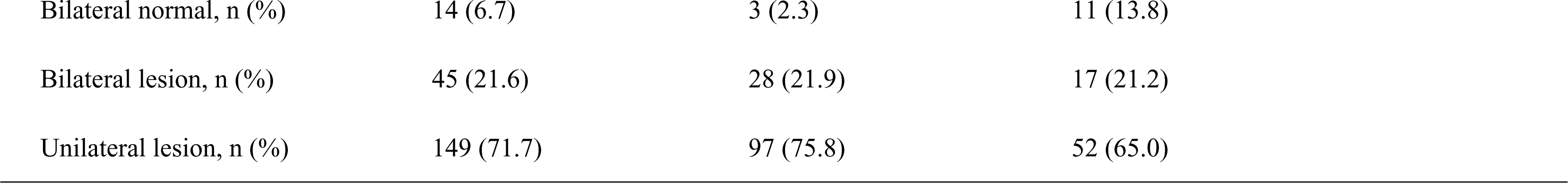
Clinical Characteristics of Patients. Data were expressed as mean ± SD, median (interquartile range) or the number (%); UPA: unilateral primary aldosteronism; BPA: bilateral primary aldosteronism; BMI: body mass index; SBP: systolic blood pressure; DBP: diastolic blood pressure; PAC: plasma aldosterone concentration; PRC: plasma renin concentration; CCT: captopril challenge test; SSIT: seated saline infusion test; * Serum potassium was the lowest level in the medical history before treatment of hypokalemia.

| Characteristic | Total (n=208) | UPA (n=128) | BPA (n=80) | P Value |
| --- | --- | --- | --- | --- |
| Age (y) | 49±11 | 49±11 | 49±10 | 0.901 |
| Female, n (%) | 97 (46.6) | 64 (50.0) | 33 (41.3) | 0.218 |
| BMI (kg/m<sup>2</sup>) | 25.5±3.4 | 25.0±3.4 | 26.3±3.2 | 0.008 |
| SBP (mm Hg) | 149 (137,163) | 147 (137,162) | 152 (137,164) | 0.544 |
| DBP (mm Hg) | 93 (85,104) | 94 (85,104) | 93 (86,106) | 0.871 |
| Serum K<sup>+</sup> (mmol/L) | 3.2±0.7 | 3.1±0.7 | 3.5±0.5 | <0.001 |
| PAC (pg/ml) | 231 (161,362) | 262 (178,405) | 200 (152,268) | <0.001 |
| PRC (μIU/ml) | 3.5 (1.3,6.7) | 2.4 (1.0,6.0) | 4.7 (2.3,8.8) | 0.003 |
| ARR (pg·ml<sup>-1</sup>/μIU· ml<sup>-1</sup>) | 69.1 (32.1,203.2) | 115 (45,278) | 42 (25,104) | <0.001 |
| PAC post-CCT (pg/ml) | 193 (143,317) | 251 (164,370) | 161 (124,198) | <0.001 |
| PAC post-SSIT (pg/ml) | 137 (98,244) | 188 (104,326) | 110 (89,144) | <0.001 |
| CT scan findings |  |  |  | 0.006 |

|  |  |  |  |
| --- | --- | --- | --- |
| Bilateral normal, n (%) | 14 (6.7) | 3 (2.3) | 11 (13.8) |
| Bilateral lesion, n (%) | 45 (21.6) | 28 (21.9) | 17 (21.2) |
| Unilateral lesion, n (%) | 149 (71.7) | 97 (75.8) | 52 (65.0) |

CT scan findings revealed normal bilateral adrenal glands in 6.7% of patients, with a significantly lower occurrence in the UPA group (2.3%) compared to the BPA group (13.8%). Bilateral lesions were found in 21.6% of patients, with similar frequencies in both the UPA (21.9%) and BPA (21.2%) groups. Unilateral lesions were more common in the UPA group (75.8%) compared to the BPA group (65.0%) (Table 1).

### 2. Correlation of parameters between ^68^Ga-Pentixafor PET/CT and AVS

Successful AVS was performed on 190 patients, with 111 diagnosed with UPA and 79 with BPA. Compared to the BPA group, UPA patients exhibited significantly higher values in the dominant side of SUV_max_ in the adrenal gland, DSAL, dominant side of PAC and aldosterone-to-cortisol ratio (ACR) (P < 0.001). The LI based on SUV_max_ in the UPA group was 1.55 times higher than in the BPA group (P < 0.001), and the LI based on AVS was 6.41 times greater in the UPA group (P < 0.001). (eTable 1)

The ACR in AVS has a significantly positive correlation with the LI based on SUV_max_ (R= 0.33, P < 0.001). In contrast, the SUV_max_ parameters do not show significant correlations with the dominant side of PAC and PCC, as indicated in (eTable 2).

### 3. Accuracy of different diagnostic criteria of ^68^Ga-Pentixafor PET/CT in the classification of PA

As illustrated in Figure 2, AUC for LI based on SUV_max_ and visual analysis in diagnosing UPA was 0.82. Both were superior to DSAL and dominant side of SUV_max_ (P < 0.001). Visual analysis achieved a sensitivity of 73% and specificity of 88% for UPA detection. The LI based on SUV_max_ cutoff of 1.50 provided the highest Youden Index of 0.59, with a sensitivity of 68% and a specificity of 91%. When the LI based on SUV_max_ cutoff was increased to 1.65, sensitivity decreased to 61%, but specificity improved to 96% (Table 2).

**Figure 2.**
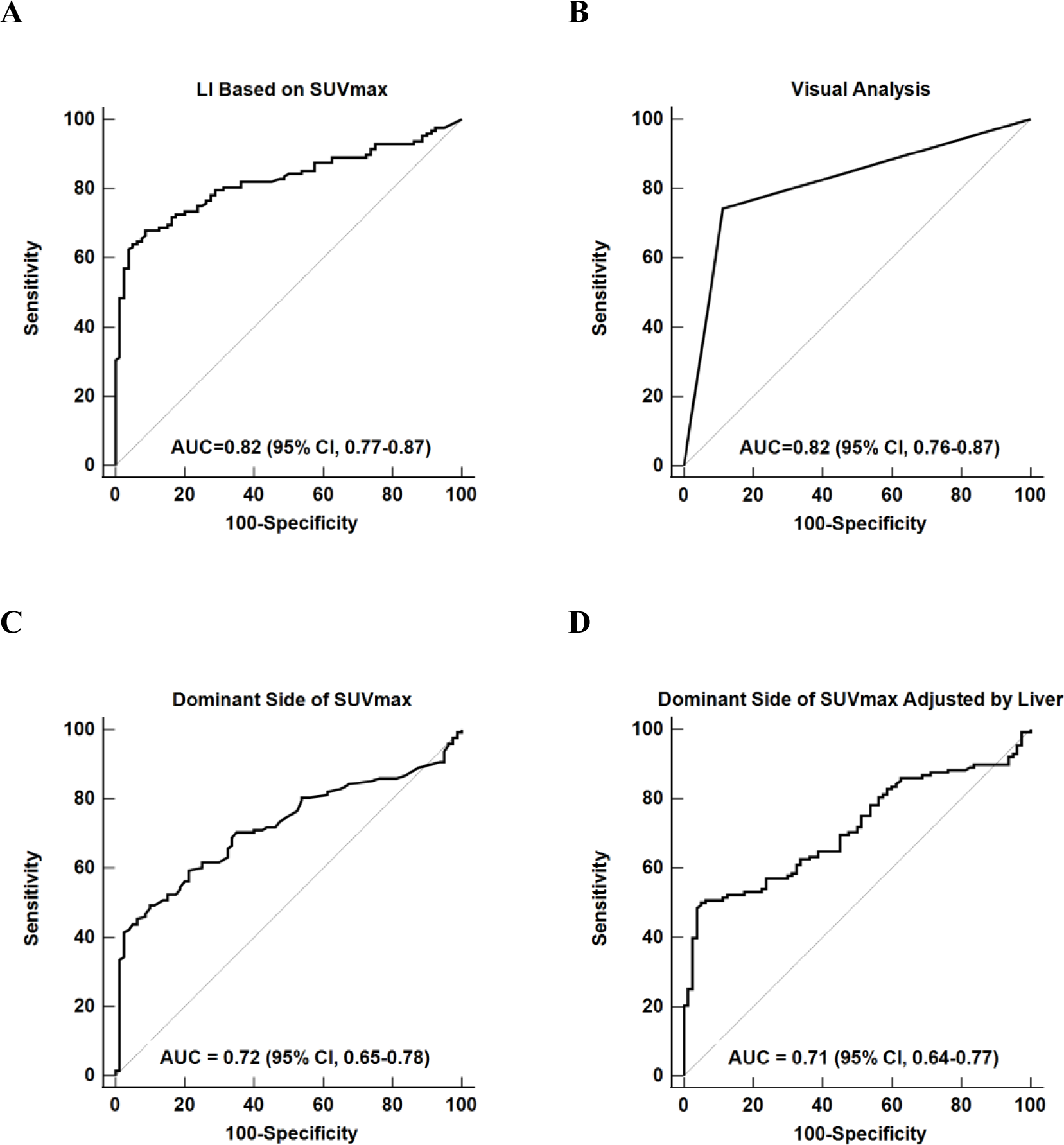
Receiver-operator Characteristic Curves of ^68^Ga-pentixafor PET/CT for the Diagnosis of Unilateral Primary Aldosteronism.

**Table 2.** Accuracy of Different Diagnostic Criteria of ^68^Ga-Pentixafor PET/CT in the Classification Diagnosis of PA. SUV_max_: maximum standardized uptake value; LI: lateralization index; TP: True Positive, FP: False Positive, FN: False Negative, TN: True Negative, PPV: positive predictive value, NPV: negative predictive value, YI: Youden index.

|  | Cutoff | TP | FP | FN | TN | Sensitivity (95% CI) | Specificity (95% CI) | YI | PPV | NPV |
| --- | --- | --- | --- | --- | --- | --- | --- | --- | --- | --- |
| LI based on SUV<sub>max</sub> | 1.35 | 95 | 19 | 33 | 61 | 0.74 (0.66-0.82) | 0.76 (0.65-0.85) | 0.50 | 0.83 (0.75-0.90) | 0.65 (0.54-0.74) |
|  | 1.50 | 87 | 7 | 41 | 73 | 0.68 (0.59-0.76) | 0.91 (0.83-0.96) | 0.59 | 0.93 (0.85-0.97) | 0.64 (0.55-0.73) |
|  | 1.65 | 79 | 3 | 49 | 77 | 0.61 (0.53-0.70) | 0.96 (0.89-1.00) | 0.58 | 0.96 (0.90-1.00) | 0.61 (0.52-0.70) |
| Visual Analysis |  | 94 | 9 | 34 | 71 | 0.73 (0.65-0.81) | 0.88 (0.80-0.95) | 0.62 | 0.91 (0.84-0.96) | 0.68 (0.58-0.76) |
| Dominant side of SUV<sub>max</sub> | 7.0 | 92 | 35 | 36 | 45 | 0.72 (0.63-0.79) | 0.56 (0.45-0.67) | 0.28 | 0.72 (0.64-0.80) | 0.56 (0.44-0.67) |
|  | 9.0 | 73 | 17 | 55 | 63 | 0.57 (0.48-0.66) | 0.79 (0.68-0.87) | 0.36 | 0.81 (0.71-0.89) | 0.53 (0.44-0.63) |
|  | 11.0 | 56 | 4 | 72 | 76 | 0.43 (0.35-0.53) | 0.95 (0.88-0.99) | 0.39 | 0.93 (0.84-0.99) | 0.51 (0.43-0.60) |
| Dominant side of SUV<sub>max</sub> adjusted by liver | 2.5 | 109 | 50 | 19 | 30 | 0.85 (0.78-0.91) | 0.38 (0.27-0.49) | 0.23 | 0.67 (0.61-0.76) | 0.61 (0.46-0.75) |
|  | 4.7 | 64 | 4 | 64 | 76 | 0.50 (0.41-0.59) | 0.95 (0.88-0.99) | 0.45 | 0.94 (0.86-0.98) | 0.54 (0.46-0.63) |
|  | 5.4 | 50 | 2 | 78 | 78 | 0.39 (0.31-0.48) | 0.98 (0.91-1.00) | 0.37 | 0.96 (0.87-1.00) | 0.50 (0.42-0.58) |

To ensure specific localization to the left or right adrenal gland rather than simply distinguishing unilateral from bilateral disease, we assessed the concordance rate between subtyping diagnosis based on ^68^Ga-Pentixafor PET/CT and diagnosis based on AVS and/or surgery (Table 3). Concordance rates for LI based on SUV_max_ cutoff points at 1.5 and 1.65 were 76.9% and 75.0%, respectively, and visual analysis had a consistency rate of 78.4%. Notably, visual analysis yielded conflicting results in the unilateral functional localization diagnosis of two patients.

**Table 3.**
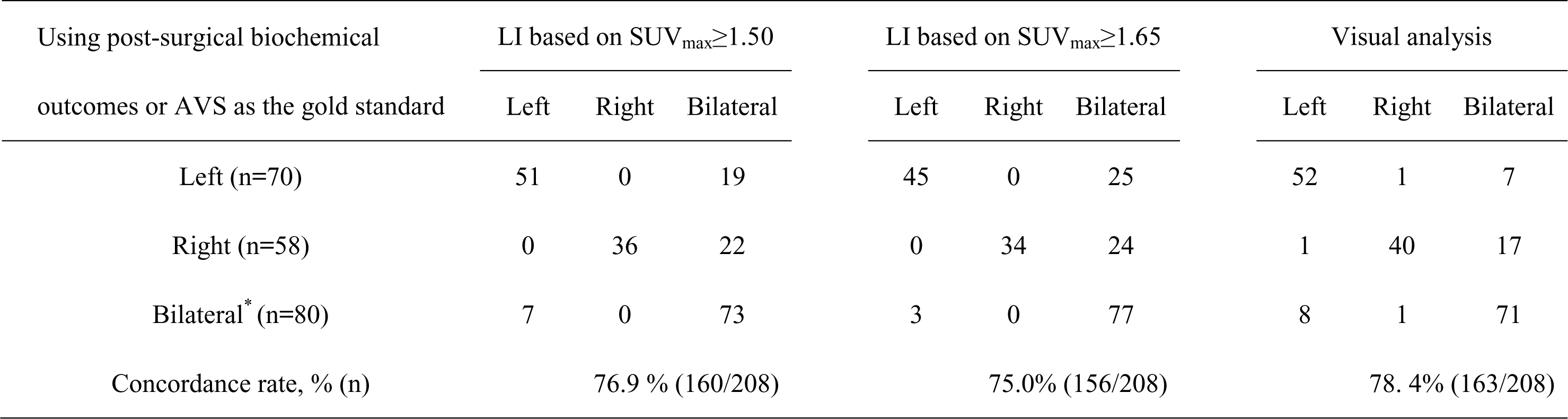
Concordance Rate Between Subtyping Diagnosis Based on ^68^Ga-Pentixafor PET/CT and Diagnosis Based on AVS and/or Surgery. *Bilateral normal or bilateral lesion.

### 4. Subgroup analysis

Patients were stratified into subgroups based on adrenal CT: those with nodules ≥10 mm (including unilateral and/or bilateral, n=112), with nodules <10 mm (43) and without nodules (including cases of hyperplasia, and normal adrenals, n=53). Subgroup analysis (eTable3) showed that the ROC for all four criteria was higher in patients with ≥10mm nodules (0.77-0.87) than that in patients with less than 10mm nodules (0.53-0.73) or without nodules (0.51-0.60). In patients with nodules ≥10mm, the ROC of LI based on SUV_max_ was higher than that of visual analysis (0.87 vs 0.77). However, the concordance rate to AVS and/or Surgery between LI based on SUV_max_ (83.9%%) and visual analysis (83.0%) was similar (eTable4). When the cutoff of LI based on SUV_max_ was 1.50, the sensitivity and specificity were 0.85(95%CI,0.76-0.91) and 0.81(95%CI,0.58-0.95) respectively. Increasing the cutoff to 1.65, the sensitivity decreased to 0.79 (95%CI,0.70-0.87), and the specificity increased to 0.90(95%CI,0.70-0.99). The sensitivity of visual analysis [0.88(95%CI,0.79-0.94)] was higher than LI based on SUV_max_, but the specificity [0.67 (95%CI,0.43-0.85)] was lower. While in those with nodules <10 mm or without nodules, both LI based on SUV_max_ and visual analysis resulted a low sensitivity of 10%-56%, with a specificity of 93%-100% (eTable 5). The smallest nodule that could be identified was 8 mm.

## Discussion

^68^Ga-Pentixafor PET/CT is an emerging method for the classification diagnosis PA, and our study including the largest number of PA patients to date. Using AVS and/or post-surgical biochemical outcomes as the reference standard, we compared the accuracy of four diagnostic criteria of ^68^Ga-Pentixafor PET/CT in differentiating UPA from BPA. Our study demonstrated that LI based on SUV_max_ and visual analysis was superior to dominant side of SUV_max_ and dominant side of SUVmax adjusted by liver, with the two criteria showing comparable diagnostic accuracy. Nevertheless, visual analysis is more sensitive, and LI based on SUV_max_ is more advantageous in specificity. Notably, this new method should be applied to PA patients with obvious nodule (diameter larger than 10mm) on CT scan, as for those with small nodule or without nodule, ^68^Ga-Pentixafor PET/CT has a high probability of missing patients with unilateral lesions who would have a better prognosis with surgery.

Heinze et al. firstly proposed using ^68^Ga-pentixafor PET/CT for diagnosing APA in 2018. They summarized the imaging results of 9 patients with APA and 44 non-PA with normal adrenal gland, using the SUV_max_ to identify APA showed a ROC of 0.964, with a cutoff value of 4.9 for SUV_max_ demonstrating a sensitivity of 88.9% and a specificity of 87.2%^13^. Later, other criteria for ^68^Ga-Pentixafor PET/CT in sub-typing PA have been reported, including the LI based on SUV_max_, liver-adjusted SUV_max_, and visual analysis. In a study by Ding et al., 25 patients with APA, 4 with idiopathic adrenal hyperplasia (IAH), and 10 with non-functional adenoma (NFA) were prospectively recruited. In the four criteria reported, a cutoff value for lesion-to-liver ratio of 2.36 reached the highest accuracy, with 100% sensitivity and 100% specificity to identify APA^12^. Similarly, in a study recruiting 26 PA patients (19 UPA and 7 BPA), among the four criteria reported, a cutoff value for lesion-to-liver ratio of 3.05 reached the highest sensitivity of 94.74% and specificity of 100% to identify UPA. The author recommended lesion-to-liver ratio as the best index for the identification of functional lesions^9^. In another study from Gao et al., immunohistochemistry of aldosterone synthase was used to identify functional nodules in postoperative adrenal tissue sections from 56 patients (37 with APA, 9 with IHA, and 10 with NFA), and 43 functional nodules and 13 non-functional nodules were included. In comparison the visual analysis with SUVmax to identify functional nodules, they found a higher accuracy of visual analysis (93.9% vs 89.8%).^10^. Zheng et al. analyzed 120 patients, diagnosing 66 with APA, 33 with IHA, and 21 with NFA. In comparison the four criteria, they proposed visual analysis as the best criteria, with sensitivity, specificity, and accuracy of 92.4%, 94.4%, and 93.33%, respectively^8^. In our previous, we compared three criteria (i.e., LI based on SUV_max_, liver-adjusted SUV_max_, and SUVmax) and we found LI based on SUV_max_ was better than liver-adjusted SUV_max_, and SUVmax. Here, we verified the superiority of LI based on SUV_max_ to the other two criteria and found LI based on SUV_max_ was comparable to visual analysis. One of the reasons for the inconsistency of the results of the above studies may be the different reference standard used for subtyping in each study. Our current and previous study mainly used AVS as the reference standard (91.3% and 100% patients had AVS, respectively)^7^. While other teams’ studies were mainly based on surgical outcomes (pathological or biochemical)^8,10-12^, and the number of BPA patients (who would be more difficult to be differentiated with UPA than the patients with NFA) included was limited.

In our study, for the patients with nodules ≥10 mm, ROC of all the criteria were higher than that in the patients with nodules <10 mm or without nodule. Two published studies have shown high accuracy of ^68^Ga-Pentixafor PET/CT in PA patients with nodules smaller than 1cm^8,11^, however, our results revealed a low sensitivity in patients with nodules <10 mm or without nodule whatever criteria used for subtyping. In our study, the smallest nodules identified by PET-CT was 8mm in diameter, which was close to 10mm, and we observed that the diameter of nodules in surgical specimens from these patients often reached or exceeded 10mm (unpublished data). In clinical practice, there could be artificial errors in CT measurement of nodule diameter, which may partly explain the inconsistency between our study and previous studies.

The strengths of the study include the rigorous protocol applied to the diagnosis of PA, the definite diagnoses of UPA as defined by surgical cure in 88.7% (94/106) or clear-cut lateralization on AVS in the remainder and evaluation of PET-CT by two independent doctors. But this study also has several limitations. Firstly, being retrospective, it might have caused selection biases. Conducted at a single center, its results may not be generalizable. Future research should include multicenter studies with standardized methods to validate and improve these findings. Second is the sample size of patients with BPA. Although this is the largest ^68^Ga-Pentixafor study in PA patients to date, the specificity was tested only in 80 patients with BPA, which needs to be further verified in more patients with BPA. In addition, 18.8% (24/128) UPA patients in our study did not undergo surgery. Although AVS is widely used for subtyping of PA, surgery and follow-up would further confirm the diagnosis of UPA.

AVS is commonly used for PA subtyping, but is costly, time-consuming, and technically complex, limiting its widespread use. While AVS remains a valuable tool for PA subtyping, ^68^Ga-Pentixafor PET/CT offers a viable alternative that can streamline the diagnostic process and reduce dependence on AVS, especially in patients with adrenal nodules measuring 10 mm or larger. As for the diagnosis criteria of ^68^Ga-Pentixafor PET/CT, LI and visual analysis of ^68^Ga-Pentixafor PET/CT showed comparable accuracy, and both criteria could be used in the classification diagnosis of PA. Nevertheless, visual analysis is more sensitive, and LI based on SUV_max_ is more advantageous in specificity.

## Perspectives

Our study evaluated the accuracy of various criteria for ^68^Ga-Pentixafor PET/CT in diagnosing of PA. We found that both LI based on SUV_max_ and visual analysis of ^68^Ga-Pentixafor PET/CT could be effective for classifying PA. However, this method should be limited in patients with nodules ≥ 10mm.Therefore, it is crucial for each center to validate these diagnostic criteria before implementation to ensure they are suitable for their specific patient population.

## Novelty and Significance

### What Is New?

^68^Ga-Pentixafor PET/CT is a novel technique for diagnosing PA. However, the optimal diagnostic criteria for using this method in PA classification remain a topic of debate. The strengths of this study include the rigorous protocol applied for the diagnosis of primary PA and the definitive diagnoses of UPA achieved through surgical cure or clear lateralization on AVS. This is the largest study to date using ^68^Ga-Pentixafor in patients with PA, assessing diagnostic indicators and cut points.

### What Is Relevant?

Our study demonstrated that both LI based on SUV_max_ and visual analysis of ^68^Ga-Pentixafor PET/CT can be used to diagnose UPA. However, this method is particularly recommended for patients with nodules larger than 10 mm.

### Clinical/Pathophysiological Implications?

As for the diagnosis criteria of ^68^Ga-Pentixafor PET/CT, both the LI based on SUV_max_ and visual demonstrated comparable accuracy. Visual analysis proved to be more sensitive, while LI based on SUV_max_ is more advantageous in specificity.

## Nonstandard Abbreviations and Acronyms

PA: Primary Aldosteronism
UPA: Unilateral Primary Aldosteronism
BPA: Bilateral Primary Aldosteronism
AVS: Adrenal Venous Sampling
ACTH: Adrenocorticotropic Hormone
LI: Lateralization Index
SI: Selectivity Index
ARR: Plasma Aldosterone/Renin Ratio
PRC: Plasma Renin Concentration
PAC: Plasma Aldosterone Concentration
SUV_max_: maximum standardized uptake value

## Acknowledgements

We thank other members of the Chongqing Primary Aldosteronism Study (CONPASS) Group: Mei Mei, MD, PhD; Suxin Luo, MD, PhD; Kangla Liao, MD; Yao Zhang, MD, PhD; Yunfeng He, MD, PhD; Yihong He, MD; Ming Xiao, PhD; and Bin Peng, PhD for suggestions of study design and revision.

## Author Contributions

Conception and design: Q.L., H.P., and S.Y.

Collection and assembly of data: H. S., J.H, A.Z., W.H., and Z. F.

Analysis and interpretation of the data: X.Z., F.H., and Y.S.

Drafting of the article: X.Z.

Critical revision of the article for important intellectual content: J.Y., J.H., H.S., A.Z., and Z. F.

Obtaining of funding: Q.L., Y. S., S.Y., and J.H.

Administrative, technical, or logistic support: J.Y., J.H., H.S., A.Z., and W.H.

## Data availability

The data that support the findings of this study are available from the corresponding author (Q.L., H.P., and S.Y.), upon reasonable request.

## All authors information

Xiangshuang Zhang: Department of Endocrinology, The First Affiliated Hospital of Chongqing Medical University, Chongqing, China.

Furong He: Department of Endocrinology, The First Affiliated Hospital of Chongqing Medical University, Chongqing, China.

Ying Song: Department of Endocrinology, The First Affiliated Hospital of Chongqing Medical University, Chongqing, China.

Ying Jing: Department of Endocrinology, The First Affiliated Hospital of Chongqing Medical University, Chongqing, China.

Jinbo Hu: Department of Endocrinology, The First Affiliated Hospital of Chongqing Medical University, Chongqing, China.

Hang Shen: Department of Endocrinology, The First Affiliated Hospital of Chongqing Medical University, Chongqing, China.

Aipin Zhang: Graduate Administration Office, The First Affiliated Hospital of Chongqing Medical University, Chongqing, China.

Wenwen He: Department of Endocrinology, The First Affiliated Hospital of Chongqing Medical University, Chongqing, China.

Zhengping Feng: Department of Endocrinology, The First Affiliated Hospital of Chongqing Medical University, Chongqing, China.

Qifu Li: Department of Endocrinology, The First Affiliated Hospital of Chongqing Medical University, Chongqing, China.

Hua Pang: Department of Nuclear medicine, The First Affiliated Hospital of Chongqing Medical University, Chongqing, China.

Shumin Yang: Department of Endocrinology, The First Affiliated Hospital of Chongqing Medical University, Chongqing, China.

## Supplemental Material

Tables S1–S5

